# mRNA Vaccine Effectiveness against COVID-19 among Symptomatic Outpatients Aged ≥16 Years in the United States, February – May 2021

**DOI:** 10.1101/2021.07.20.21260647

**Authors:** Sara S Kim, Jessie R Chung, Edward A Belongia, Huong Q McLean, Jennifer P King, Mary Patricia Nowalk, Richard K Zimmerman, Goundappa K Balasubramani, Emily T Martin, Arnold S Monto, Lois E Lamerato, Manjusha Gaglani, Michael E Smith, Kayan M Dunnigan, Michael L Jackson, Lisa A Jackson, Mark W Tenforde, Jennifer R Verani, Miwako Kobayashi, Stephanie Schrag, Manish M Patel, Brendan Flannery

## Abstract

Evaluations of vaccine effectiveness (VE) are important to monitor as COVID-19 vaccines are introduced in the general population. Research staff enrolled symptomatic participants seeking outpatient medical care for COVID-19-like illness or SARS-CoV-2 testing from a multisite network. VE was evaluated using the test-negative design. Among 236 SARS-CoV-2 nucleic acid amplification test-positive and 576 test-negative participants aged ≥16 years, VE of mRNA vaccines against COVID-19 was 91% (95% CI: 83-95) for full vaccination and 75% (95% CI: 55-87) for partial vaccination. Vaccination was associated with prevention of most COVID-19 cases among people seeking outpatient medical care.

## Background

Randomized controlled trials and real-world effectiveness studies have demonstrated high COVID-19 vaccine effectiveness (VE) against severe outcomes and symptomatic illness among priority groups for vaccination, including health care workers and persons aged ≥65 years [1-5]. Following the Advisory Committee on Immunization Practice’s (ACIP) recommendations for COVID-19 vaccine allocation to target populations, states expanded vaccine availability to the general public aged ≥16 years starting in the spring of 2021 [6]. Given the more common clinical presentation of mild to moderate illness compared to severe outcomes, data on VE for the prevention of COVID-19 among persons seeking care for COVID-19-like illness (CLI) in outpatient settings are needed [7].

Since 2008, the United States Influenza Vaccine Effectiveness Network (U.S. Flu VE Network) has provided influenza VE estimates annually. The strength of this long-standing active surveillance network includes coupling of clinical and epidemiological data in thousands of patients annually to generate VE estimates mid-way through each influenza season. These estimates provide decision-makers with real-time data to assess VE in the current season and contribute to informing global annual vaccine strain selection decisions. Investigations of VE in outpatient settings can enhance our understanding of protection among persons seeking care for mild or moderate illness, contribute to estimating averted healthcare burden attributed to COVID-19, and inform community mitigation policies as vaccine coverage continues to increase among adults and adolescents in the United States (U.S.). We used the robust surveillance platform of the U.S. Flu VE Network to estimate VE against laboratory-confirmed SARS-CoV-2 infection among persons aged ≥16 years with COVID-19-like symptoms seeking outpatient care or clinical SARS-CoV-2 testing.

## Methods

We used the test-negative design to evaluate messenger RNA (mRNA) VE against outpatient COVID-19 by comparing vaccine receipt in persons testing positive and negative for SARS-CoV-2 infection [8]. Beginning in March 2020, participating institutions at five study sites for the U.S. Flu VE Network in Michigan, Pennsylvania, Texas, Washington, and Wisconsin began active surveillance for COVID-19.

Research staff screened persons who sought outpatient medical care (i.e., telehealth, primary care, urgent care, and emergency departments) or clinical SARS-CoV-2 testing using a standard case-definition for CLI of an acute onset of fever/feverishness, cough, or loss of taste or smell with symptom duration <10 days [9]. Research staff contacted potentially eligible outpatients by telephone or electronic message to confirm eligibility and enroll consenting participants. In addition to meeting the CLI definition, eligible participants had a clinical or research respiratory specimen collected for SARS-CoV-2 molecular testing within 10 days of illness onset. Standardized questionnaires collected demographic information, general health status, self-reported COVID-19 vaccination, and history of individual respiratory, gastrointestinal and systemic symptoms experienced during acute illness, as well as potential risk factors for contracting COVID-19 such as working in a healthcare setting and having contact with a person with laboratory-confirmed COVID-19. SARS-CoV-2 nucleic acid amplification test results were used to classify SARS-CoV-2 positive cases and test-negative controls. Research testing, or testing for the purpose of this study, was performed if clinical results were unavailable for study use.

For this analysis, we included participants with illness onset on or after February 1, 2021, for those aged ≥65 years and on or after March 22, 2021, for those aged 16-64 years; beginning dates of inclusion in analyses varied by site according to local COVID-19 vaccination policies for all persons aged ≥65 or ≥16 years (Supplemental Table 1). We determined vaccination status through participant interviews and verified vaccination based on participant-provided vaccination record cards, documentation of vaccination in electronic medical records or state immunization information systems. Fully vaccinated participants were defined as those who received 2 doses of an mRNA vaccine (Pfizer-BioNTech BNT162b2 or Moderna mRNA-1273) ≥14 days before illness onset [2,4]. Partially vaccinated participants were defined as those who received at least one dose of an mRNA vaccine ≥14 days before illness onset but who were not fully vaccinated. Those who did not report vaccine receipt and had no documentation of any COVID-19 vaccine prior to illness were defined as unvaccinated. Participants who received their first dose <14 days prior to illness (n=100), vaccinated with Johnson and Johnson’s Janssen (JNJ-784367350) vaccine (n=22), or self-reported vaccine receipt without documentation (n=35) were excluded.

For each category of COVID-19 vaccination, VE was calculated as 1 – odds ratio of vaccination among SARS-CoV-2 test-positive participants versus test-negative participants (controls) using multivariable logistic regression. Models were adjusted *a priori* for study site, age in years (continuous), and enrollment period (natural cubic spline with 3 percentile knots of interval between January 1^st^, 2021, and illness onset date). We evaluated sex, race and Hispanic ethnicity, and having had a SARS-CoV-2 positive contact as additional covariates and included race/ethnicity and positive contact in the final models. We also performed sensitivity analyses comparing VE using plausible self-report with documented vaccination, where plausibility was determined by ability to report credible location of vaccination. Statistical analyses were conducted using SAS version 9.4. This activity was reviewed by the institutional review boards of the Centers for Disease Control and Prevention (CDC) and other participating institutions and was conducted consistent with applicable federal law and CDC policy.

## Results

Between February 1 and May 28, 2021, 27% of outpatients who were contacted for screening and enrollment agreed to participate. Among 812 enrolled participants aged ≥16 years with CLI, 236 (29%) tested positive for SARS-CoV-2. SARS-CoV-2 positivity was higher among males, participants identifying as non-Hispanic Black, subjects aged <65 years, and the Michigan and Pennsylvania study sites (Table 1). Within the enrollment period, SARS-CoV-2 positivity peaked during the second week of April.

**Table 1:**
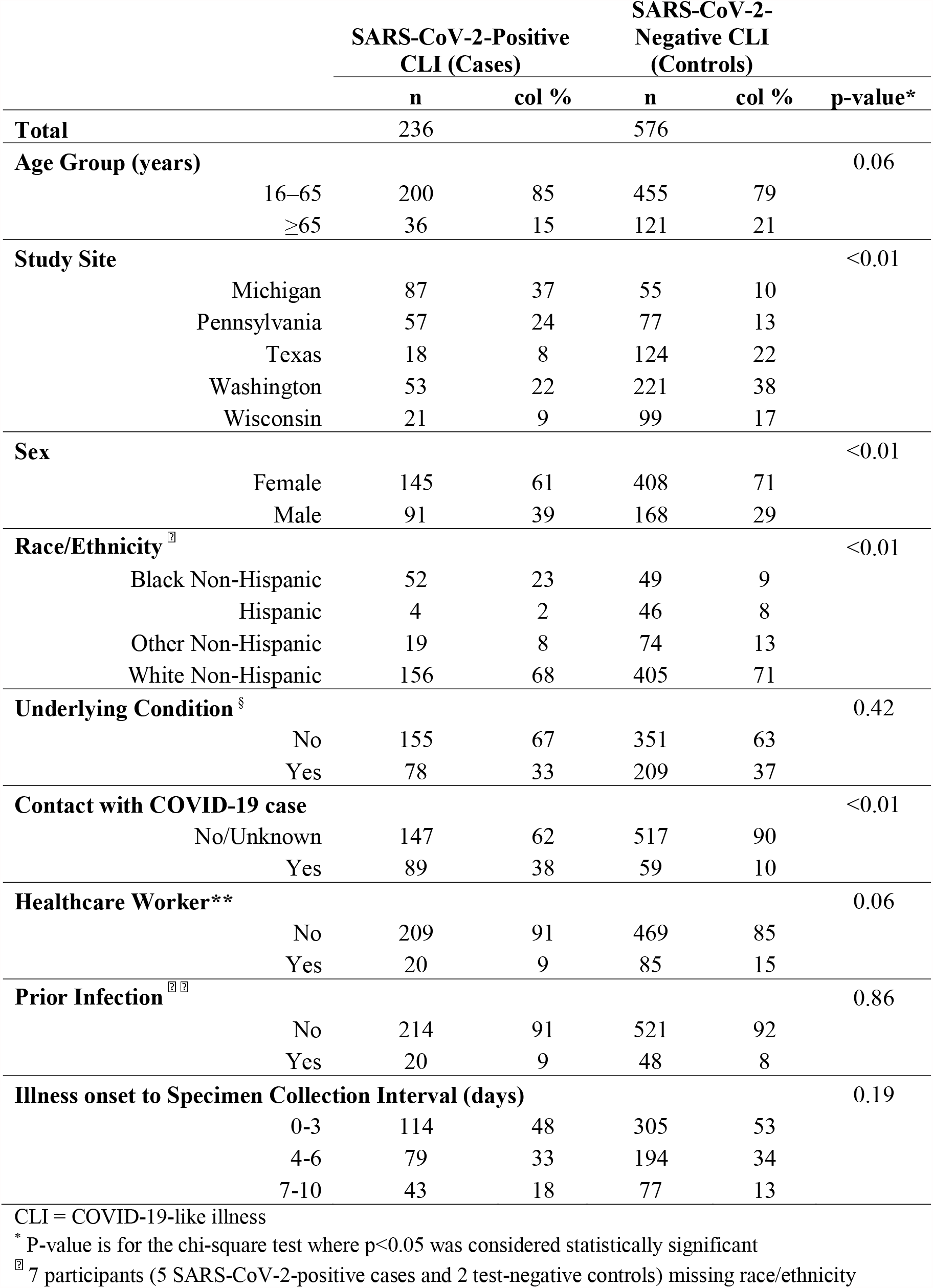
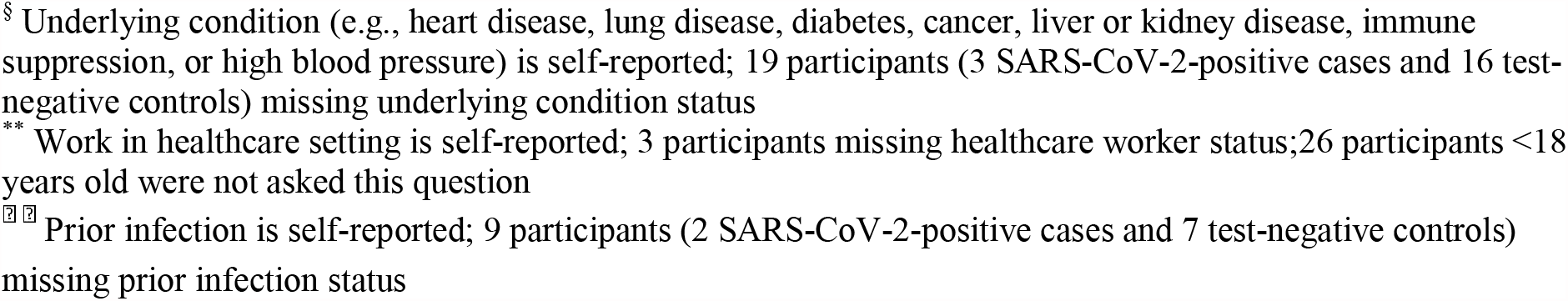
Characteristics of Enrolled Participants by SARS-CoV-2 Status, U.S. Flu VE Network, February 1-May 28, 2021

|  | SARS-CoV-2-Positive<br>CLI (Cases) |  | SARS-CoV-2-Negative CLI<br>(Controls) |  | p-value* |
| --- | --- | --- | --- | --- | --- |
|  | n | col % | n | col % |  |
| **Total** | 236 |  | 576 |  |  |
| **Age Group (years)** |  |  |  |  | 0.06 |
| 16–65 | 200 | 85 | 455 | 79 |  |
| ≥65 | 36 | 15 | 121 | 21 |  |
| **Study Site** |  |  |  |  | <0.01 |
| Michigan | 87 | 37 | 55 | 10 |  |
| Pennsylvania | 57 | 24 | 77 | 13 |  |
| Texas | 18 | 8 | 124 | 22 |  |
| Washington | 53 | 22 | 221 | 38 |  |
| Wisconsin | 21 | 9 | 99 | 17 |  |
| **Sex** |  |  |  |  | <0.01 |
| Female | 145 | 61 | 408 | 71 |  |
| Male | 91 | 39 | 168 | 29 |  |
| **Race/Ethnicity<sup>‡</sup>** |  |  |  |  | <0.01 |
| Black Non-Hispanic | 52 | 23 | 49 | 9 |  |
| Hispanic | 4 | 2 | 46 | 8 |  |
| Other Non-Hispanic | 19 | 8 | 74 | 13 |  |
| White Non-Hispanic | 156 | 68 | 405 | 71 |  |
| **Underlying Condition<sup>§</sup>** |  |  |  |  | 0.42 |
| No | 155 | 67 | 351 | 63 |  |
| Yes | 78 | 33 | 209 | 37 |  |
| **Contact with COVID-19 case** |  |  |  |  | <0.01 |
| No/Unknown | 147 | 62 | 517 | 90 |  |
| Yes | 89 | 38 | 59 | 10 |  |
| **Healthcare Worker\*\*** |  |  |  |  | 0.06 |
| No | 209 | 91 | 469 | 85 |  |
| Yes | 20 | 9 | 85 | 15 |  |
| **Prior Infection<sup>‡ ‡</sup>** |  |  |  |  | 0.86 |
| No | 214 | 91 | 521 | 92 |  |
| Yes | 20 | 9 | 48 | 8 |  |
| **Illness onset to Specimen Collection Interval (days)** |  |  |  |  | 0.19 |
| 0-3 | 114 | 48 | 305 | 53 |  |
| 4-6 | 79 | 33 | 194 | 34 |  |
| 7-10 | 43 | 18 | 77 | 13 |  |

CLI = COVID-19-like illness
\* P-value is for the chi-square test where p<0.05 was considered statistically significant
<sup>‡</sup> 7 participants (5 SARS-CoV-2-positive cases and 2 test-negative controls) missing race/ethnicity
<sup>§</sup> Underlying condition (e.g., heart disease, lung disease, diabetes, cancer, liver or kidney disease, immune suppression, or high blood pressure) is self-reported; 19 participants (3 SARS-CoV-2-positive cases and 16 test-negative controls) missing underlying condition status
<sup>\*\*</sup> Work in healthcare setting is self-reported; 3 participants missing healthcare worker status; 26 participants <18 years old were not asked this question
<sup>¶</sup> <sup>¶</sup> Prior infection is self-reported; 9 participants (2 SARS-CoV-2-positive cases and 7 test-negative controls) missing prior infection status

Across all vaccinated participants included in the analysis, 226 (62%) received Pfizer-BioNTech, and 138 (38%) received the Moderna vaccine. Among vaccinated SARS-CoV-2 positive cases, 37 (16%) received at least one dose of an mRNA COVID-19 vaccine. Seventeen (46%) of the 37 cases who received any vaccine dose were considered fully vaccinated, of which 15 received Pfizer-BioNTech and 2 received Moderna; 20 (54%) of the 37 cases were partially vaccinated, of which 15 received Pfizer-BioNTech and 5 received Moderna. In comparison, 327 (57%) SARS-CoV-2 negative controls were vaccinated with at least one dose, of which 231 (71%) were fully vaccinated and 96 (29%) were partially vaccinated.

Effectiveness of mRNA vaccines against laboratory-confirmed COVID-19 in outpatient settings was 91% (95% Confidence Interval [CI]: 83-95%) among those fully vaccinated and 75% (95% CI: 55-87%) among those partially vaccinated (Table 2). VE was similar when using documentation, plausible self-report, or both to classify vaccination status (Supplemental Table 2).

**Table 2:** Estimates of mRNA Vaccine Effectiveness against Laboratory-Confirmed COVID-19 among Outpatients, Using Vaccine Doses Verified by Immunization Documentation

|  | SARS-CoV-2-Positive CLI<br>(Cases) |  |  | SARS-CoV-2-Negative CLI<br>Controls |  |  | Unadjusted VE |  | Adjusted* VE |  |
| --- | --- | --- | --- | --- | --- | --- | --- | --- | --- | --- |
|  | # |  | % | # |  | % | VE |  |  |  |
|  | Vaccinated | Total | Vaccinated | Vaccinated | Total | Vaccinated | % | (95% CI) | VE % | (95% CI) |
| **Vaccination Status** |  |  |  |  |  |  |  |  |  |  |
| Full vaccination | 17 | 216 | 8 | 231 | 480 | 48 | 91 | (84-95) | 91 | (83-95) |
| Partial vaccination | 20 | 219 | 9 | 96 | 345 | 28 | 74 | (56-84) | 75 | (55-87) |

CLI = COVID-19-like illness
\*Vaccine effectiveness (VE) adjusted for study site, age in years (continuous), enrollment period (natural cubic spline with 3 percentile knots of interval between January 1, 2021 and illness onset date), race/ethnicity, and contact with a SARS-CoV-2 case

## Discussion

During February–May 2021 in a multisite outpatient network evaluating COVID-19 vaccine effectiveness, vaccination reduced laboratory-confirmed symptomatic illness by 91% among those fully vaccinated and 75% among those partially vaccinated. These findings add to evidence from clinical trials of efficacy against symptomatic illness and from observational studies of vaccine effectiveness across the continuum of illness severity in multiple countries [1-5,10-12].

Monitoring vaccine effectiveness against COVID-19 in outpatient settings is relevant for three reasons. First, outpatient settings may better capture younger age groups that account for an increasing proportion of COVID-19 cases and are likely to present with moderate symptoms necessitating outpatient care rather than hospitalization [13]. Furthermore, vaccine coverage is lower in younger adults and adolescents, and thus a higher proportion of cases may occur in this age group. Second, people with mild and moderate COVID-19 are rarely hospitalized and more likely to seek care in outpatient facilities. Thus, when considering logistics of monitoring vaccine effectiveness, planning for enrollment and sample size, evaluating duration of protection, and assessing protection against variants of concern in real-time is more feasible in outpatient settings. Third, past experiences from the annual influenza vaccination program have demonstrated success of these types of studies for monitoring vaccine effectiveness among people seeking outpatient medical care for respiratory illness. In turn, vaccine effectiveness studies in outpatient settings can be conducted in a timely manner and have less potential for confounding [8]. Thus, implementing VE studies in outpatient settings to monitor vaccine protection could be valuable for informing policy decisions such as community mitigation strategies and strain selection for vaccines or booster doses.

These results were subject to several limitations. First, because Johnson & Johnson’s Janssen vaccine uptake in the general population was limited during the study period, participants vaccinated with the Janssen vaccine were not included in this analysis. Including the 22 vaccinated participants who received Janssen vaccine (4 in cases, 18 in controls) resulted in similar estimates of VE (Supplemental Table 2). Second, surveillance populations at the study sites are not representative of the U.S. population, and this analysis did not evaluate VE by race and Hispanic ethnicity. Additional evaluation of VE among racial/ethnic groups disproportionately affected by the pandemic and among other specific populations, such persons with underlying health conditions, are needed. Third, because we relied on vaccine documentation to determine vaccination status, we may have missed unrecorded vaccine doses. However, estimates including self-reported doses, without documentation, showed similar VE (Supplemental Table 2). Finally, unmeasured selection bias based on healthcare seeking behaviors and potential for differential participation response is a possible risk in the test-negative design.

As of July 1, 2021, 55% of the U.S. population had received at least one dose of a COVID-19 vaccine [14]. A growing number of vaccine effectiveness studies have provided evidence that mRNA vaccines confer similar protection against COVID-19 in real-world conditions as in clinical trials, reducing risk of infection and related severe outcomes by 90% or more among those fully vaccinated [1,2,11]. In this study, receipt of mRNA vaccines was associated with prevention of most mild to moderate COVID-19 in outpatients seeking medical care or testing in the U.S.

Studies should continue to monitor COVID-19 vaccine effectiveness over time and against variant SARS-CoV-2 viruses to inform vaccination strategies. The findings of this investigation support ACIP recommendations to vaccinate eligible persons as well as efforts to increase vaccine coverage in the U.S. population for the prevention of symptomatic illness as restrictions regarding mask wearing, social distancing, and school and workplace closures change. These data suggest that continued efforts to vaccinate U.S. adults and adolescents are warranted and will likely have a marked impact on mild to moderate COVID-19.

## Data Availability

De-identified dataset can be made available upon request

## Acknowledgements

Hannah Berger, Joshua Blake, Keegan Brighton, Gina Burbey, Deanna Cole, Linda Heeren, Erin Higdon, Lynn Ivacic, Julie Karl, Sarah Kopitzke, Erik Kronholm, Jennifer Meece, Nidhi Mehta, Vicki Moon, Cory Pike, Carla Rottscheit, Jackie Salzwedel, Marshfield Clinic Research Institute, Marshfield, Wisconsin; Alanna Peterson, Linda Haynes, Erin Bowser, Louise Taylor, Karen Clarke, Krissy Moehling Geffel, Todd M. Bear, Klancie Dauer, Heather Eng, Monika Johnson, Donald B. Middleton, Jonathan M. Raviotta, Theresa Sax, Miles Stiegler, Joe Suyama, Alexandra Weissman, John V. Williams, University of Pittsburgh Schools of the Health Sciences and University of Pittsburgh Medical Center, Pittsburgh, Pennsylvania; Adam Lauring, Joshua G. Petrie, Lois E. Lamerato, E.J. McSpadden, Caroline K.Cheng, Rachel Truscon, Samantha Harrison, Armanda Kimberly, Anne Kaniclides, Kim Beney, Sarah Bauer, Michelle Groesbeck, Joelle Baxter, Rebecca Fong, Drew Edwards, Weronika Damek Valvano, Micah Wildes, Regina Lehmann-Wandell, Caitlyn Fisher, Luis Gago, Marco Ciavaglia, Kristen Henson, Kim Jermanus, Alexis Paul, University of Michigan, Ann Arbor, and Henry Ford Health System, Detroit, Michigan; Eric Hoffman, Martha Zayed, Marcus Volz, Kimberly Walker, Arundhati Rao, Manohar Mutnal, Michael Reis, Lydia Requenez, Amanda McKillop, Spencer Rose, Kempapura Murthy, Chandni Raiyani, Natalie Settele, Jason Ettlinger, Courtney Shaver, Elisa Priest, Jennifer Thomas, Alejandro Arroliga, Madhava Beeram, Baylor Scott & White Health, Temple Texas; C. Hallie Phillips, Erika Kiniry, Stacie Wellwood, Brianna Wickersham, Matt Nguyen, Rachael Burganowski, Suzie Park, Kaiser Permanente Washington Research Institute, Seattle, Washington.

