## Supplemental Tables for "mRNA Vaccine Effectiveness against COVID-19 among Symptomatic Outpatients Aged ≥16 Years in the United States, February – May 2021"

| **Vaccine Group Phase** | **Michigan**^*^ | **Pennsylvania** ^ᵻ^ | **Texas**^§^ | **Washington**^**^ | **Wisconsin** ^ᵻ ᵻ^ |
| --- | --- | --- | --- | --- | --- |
| **1A** | 12/2020: healthcare workers, long-term care facility residents | 12/2020: healthcare workers, long-term care facility residents, persons aged ≥65 years, persons aged 16-64 years with high-risk condition | 12/14/2020: healthcare workers, long-term care facility residents | 12/2020: healthcare workers, long-term care facility residents | 12/2020: healthcare workers, long-term care facility residents |
| **1B** | 1/2021: persons aged ≥65 years, essential frontline workers | 3/2021: essential frontline workers | 12/29/2020: persons aged ≥65 years, persons aged 16-64 years with high-risk condition;  3/3/2021: school and licensed childcare personnel | 1/18/2021: persons aged ≥65 years, persons aged ≥50 years living in multi-generational household, educators, childcare providers; 3/17/2021: essential frontline workers, pregnant individuals, persons with disability | 1/25/2021: essential frontline workers, persons aged ≥65 years |
| **1C** | 3/8/2021: persons aged 50-64 with disabilities or high-risk conditions, caregivers of children with special needs; 3/22/2021: persons aged 16-49 years with disabilities or high-risk conditions | 4/12/2021: other essential workers | 3/15/2021: persons aged 50-64 years |  | 3/22/2021: persons aged 16-64 years with high-risk condition |
| **2** | 3/22/2021: persons aged 50-64 years  4/5/2021: persons aged ≥16 years | 4/13/2021: persons aged ≥16 years | 3/29/2021: persons aged ≥16 years | 4/15/2021: persons aged ≥16 years | 4/5/2021: persons aged ≥16 years |

**Supplemental Table 1: Vaccine Group Phases by State COVID-19 Vaccination Policies**

^**^ https://www.michigan.gov/documents/coronavirus/MI_COVID-19_Vaccination_Prioritization_Guidance_2152021_716344_7.pdf

^ᵻ^ https://www.health.pa.gov/topics/disease/coronavirus/Vaccine/Pages/Distribution.aspx

^§^ https://www.dshs.state.tx.us/coronavirus/immunize/vaccineallocations.aspx

^**^ https://www.doh.wa.gov/Portals/1/Documents/1600/coronavirus/VaccinationPhasesInfographic.pdf

^ᵻ ᵻ^ https://projects.jsonline.com/topics/coronavirus/vaccine-faq/wisconsin-covid-vaccine-who-eligible-where-get.html

**Supplemental Table 2**: Sensitivity Analysis of Vaccine Effectiveness with Plausible Self-Report (PSR) and Inclusion of Johnson & Johnson’s Janssen Vaccine

|  | **SARS-CoV-2-Positive CLI (Cases)** | | | **SARS-CoV-2-Negative CLI (Controls)** | | | **Unadjusted VE** | | **Adjusted* VE** | |
| --- | --- | --- | --- | --- | --- | --- | --- | --- | --- | --- |
|  | # Vaccinated | Total | % Vaccinated | # Vaccinated | Total | % Vaccinated | VE % | (95% CI) | VE % | (95% CI) |
| **Full vaccination** |  |  |  |  |  |  |  |  |  |  |
| Documented ^ᵻ^ | 21 | 229 | 9 | 249 | 524 | 48 | 89 | (82-93) | 88 | (79-94) |
| Documented+PSR^§^ | 22 | 226 | 10 | 254 | 513 | 50 | 89 | (82-93) | 88 | (78-93) |
| PSR only | 17 | 217 | 8 | 197 | 464 | 42 | 88 | (80-93) | 89 | (78-94) |
| **Partial vaccination** |  |  |  |  |  |  |  |  |  |  |
| Documented ^ᵻ^ | 20 | 228 | 9 | 96 | 371 | 26 | 72 | (54-84) | 74 | (53-86) |
| Documented+PSR^§^ | 20 | 224 | 9 | 99 | 358 | 28 | 74 | (57-85) | 75 | (55-86) |
| PSR only | 27 | 227 | 12 | 103 | 370 | 28 | 65 | (44-78) | 65 | (38-80) |
| **Including Janssen** |  |  |  |  |  |  |  |  |  |  |
| Full Vaccination | 21 | 220 | 10 | 249 | 498 | 50 | 89 | (83-93) | 89 | (80-94) |

CLI = COVID-19-like illness

*Vaccine effectiveness (VE) adjusted for study site, age in years (continuous), enrollment period (natural cubic spline with 3 percentile knots of interval between January 1, 2021 and illness onset date), race/ethnicity, and contact with a SARS-CoV-2 case

^ᵻ^ Vaccine status based on verified doses; Persons who self-reported vaccination that could not be confirmed by documentation were excluded in the main analysis and considered unvaccinated in this sensitivity analysis

^§^ Vaccine status based on verified doses and supplementing vaccine receipt with plausible self-report in the absence of documentation
